# Gender inequalities in trajectories of depressive symptoms among young people in London and Tokyo: a longitudinal cross-cohort study

**DOI:** 10.1101/2023.11.22.23298823

**Authors:** Gemma Knowles, Daniel Stanyon, Syudo Yamasaki, Mitsuhiro Miyashita, Charlotte Gayer-Anderson, Kaori Endo, Satoshi Usami, Junko Niimura, Naomi Nakajima, Kaori Baba, TTC Young Persons Advisory Group, Thai-sha Richards, Jonas Kitisu, Adna Hashi, Karima Shyan Clement-Gbede, Niiokani Tettey, Samantha Davis, Katie Lowis, Verity Buckley, Dario Moreno-Agostino, Esther Putzgruber, Holly Crudgington, Charlotte Woodhead, Kristi Sawyer, Katherine M. Keyes, Jacqui Dyer, Shuntaro Ando, Kiyoto Kasai, Mariko Hiraiwa-Hasegawa, Craig Morgan, Atsushi Nishida

## Abstract

**Background:** Research suggests gender inequalities in adolescent mental health are context dependent. This implies they may be preventable through social/structural change. However, there is also some evidence that gender mental health gaps are *larger* in ostensibly more gender equal societies, e.g., 2-3-fold larger in the UK vs. Japan. Using data and methods that overcome important limitations of existing evidence, we tested the hypothesis that gender inequalities in depressive symptom trajectories are larger in London than in Tokyo, and that these differences are not due to incomparable measurement.

**Methods:** We used three waves of data from representative adolescent cohorts in Tokyo (TTC; *n*=2,813) and London (REACH; *n*=4,287) (*n=*7,100; age 11-16y). We used multigroup and longitudinal confirmatory factor analysis to examine measurement invariance of the 13-item Short Mood and Feelings Questionnaire (SMFQ) across cohorts, genders, and ages. Latent growth models compared depressive symptom trajectories of boys and girls in London and Tokyo.

**Outcomes:** Scalar invariance was well-supported. In London, gender inequalities in depressive symptoms were evident at age 11y (girls: +0·8 [95% CI: 0·3-1·2]); in Tokyo, the difference emerged between 11-14y. In both places, the disparity widened year-on-year, but by age 16y was around twice as large in London. Annual rate of increase in depressive symptoms was around four times steeper among girls in London (1·1 [0·9-1·3]) vs. girls in Tokyo (0·3 [0·2-0·4]).

**Interpretation:** Gender inequalities in emotional health are context dependent and may be preventable through social/structural change.

**Funding:** Japanese Society for the Promotion of Science; Economic and Social Research Council.

**RESEARCH IN CONTEXT:** *Evidence before this study:* Women are around two-to-four times more likely than men to experience emotional problems such as depression and anxiety. Our understanding of the causes is surprisingly limited. Typically, these gender inequalities in emotional health emerge in early adolescence, at around the age of puberty, so much research has focussed on biological explanations. However, a growing body of evidence suggests gender inequalities in adolescent mental health may be context dependent, varying in size – and sometimes direction – across countries. This implies it may be possible to prevent the excess of mental distress among teenage girls through social/structural change. However, there is also some evidence to suggest that gender inequalities in teenage mental health are *larger*, on average, in countries with higher levels of societal gender equity, e.g., around 2-3 times larger in the UK (which ranks 15^th^ on global gender equity) compared with Japan (ranked 125^th^). Reasons for this seemingly paradoxical trend are unclear. However, there are important limitations to the international evidence that preclude robust inference about the contexts and conditions that give rise to (and those that mitigate and prevent) gender inequalities in emotional health. It is mostly cross-sectional, relates to older age groups, or – importantly – fails or is unable to robustly examine measurement invariance between countries. We reviewed the reference lists in two successive reviews (published in 2000 and 2017) on the causes of gender inequalities in depression and searched PubMed for original and review articles published as of January 2023. Search terms included: gender inequalities (sex/gender differences, inequalities, disparities, etc.) AND mental health (mental distress, depression, depressive symptoms, etc.) AND young people (child*, adolesc*, youth, etc.) AND international comparisons (international comparisons, cross-cohort, cross-cultural, etc.). We screened titles and abstracts to identify studies with longitudinal data on mental health in population-based adolescent samples. We found: one cross-sectional study reporting gender inequalities in mental distress and wellbeing at age 15 years in 73 countries, with measurement invariance considered at the regional level (e.g., Americas, Eastern Mediterranean); one cross-sectional study of all age groups (except children under 12) in 90 countries, with no examination of measurement invariance; and four longitudinal studies comparing gender inequalities in mental health across countries in mid-adolescence, that either (a) used unrepresentative samples, (b) compared countries with very similar levels of societal gender equity, or (c) did not examine – or only partly supported – measurement invariance between countries.

*Added value of this study:* We used three waves of data from large, representative cohorts of young people in Tokyo and London and examined (a) the extent to which a widely used measure of depressive symptoms is invariant (comparable) across place, gender, and age, and (b) whether inequalities in depressive symptom trajectories between adolescent boys and girls are larger in London than in Tokyo. We found strong evidence that inequalities in depressive symptom trajectories between adolescent boys and girls are around twice as large, and may emerge earlier, among young people in London than in Tokyo. Notably, the annual rate of increase in depressive symptoms from age 11 to age 16 was around four times steeper among girls in London than among girls in Tokyo. Importantly, we found little evidence to suggest these differences are due to incomparable measurement. We co-wrote this paper with ten young people, five in London and five in Tokyo, and their perspectives are integrated throughout and presented in the Supplement.

*Implications of all the available evidence:* There is strong evidence that the size and course of gender inequalities in emotional health are driven by social/structural context. Against a backdrop of high and rising rates of emotional health problems among young women and girls in many countries, there is an urgent need to understand the contexts and conditions that enable young girls to thrive.

## INTRODUCTION

For decades, research in high-income Western countries has reported marked gender inequalities in prevalence and incidence of mental health problems.^1^ Emotional problems, including depression and anxiety, are around two-to-four times more common among women than among men.^1,2^ These inequalities typically emerge in early adolescence, peak in late adolescence, and persist across the lifecourse.^1–5^ Our understanding of the causes is limited. In adults, there is growing evidence for a role of structural gender inequity (e.g., unequal pay)^6^ and, increasingly, indications that gender inequalities in mental health are context dependent (e.g., vary by country).^7^ This implies it may be possible to reduce or prevent them with social and structural change.

However, the ways in which structural gender inequity manifests in adulthood (i.e., in unequal income, employment, childcare, etc.) are less clearly relevant to early adolescence, when gender inequalities in emotional health typically emerge. That said, emerging evidence does suggest a role of gendered violence (e.g., sexual harassment) in mid-adolescence,^8^ and there are indeed indications that, as in adults, the size – and possibly direction – of gender inequalities in mental distress in mid-adolescence varies substantively between countries.^9^ Counter-intuitively, this same evidence suggests gender inequalities in mental distress in mid-adolescence are *larger* in countries that tend to perform *better* on indices of societal gender equity.^9^ This raises interesting questions about the conditions that cause and exacerbate, and those that reduce or prevent, the onset of gender inequalities in mental health. However, there are important limitations to the existing international evidence that must first be addressed. It is largely cross-sectional,^5,9^ focusses on mid-to-late adolescence,^5,9^ uses unrepresentative samples, ^10^ compares countries that rank very similarly on societal gender equity^4,11^ or, crucially, does not consider or only partially supports measurement equivalence between countries.^5,9–12^ As such, we cannot rule out that variations in the size of gender inequalities in mental health across diverse cultural contexts are due to incomparable measurement.

In this study, we sought to address these limitations using three waves of data from two representative adolescent cohort studies in Tokyo (Japan) and London (UK): the Tokyo Teen Cohort (TTC)^13^ and Resilience, Ethnicity and AdolesCent Mental Health (REACH).^14^ At the nation-level, the UK and Japan have similarly high levels of economic development but starkly different scores on gender equity. For instance, the Global Gender Gap Index (GGGI) ranks the UK 15^th^ out of 149 countries while Japan ranks 15^th^.^15^ We aimed to (a) examine the extent to which a widely used measure of depressive symptoms is invariant across cohorts, genders, and age, and (b) compare gender inequalities in depressive symptom trajectories from age 11 to 16 years. Specifically, we tested the hypotheses that inequalities in depressive symptom trajectories between boys and girls are larger in London than in Tokyo, and that these differences are not due to incomparable measurement.

## METHODS

### Study design and participants

TTC is a population-based cohort study of adolescent health and development based in three districts of Tokyo (Setagaya, Mitaka, Chofu) (http://ttcp.umin.jp/).^13^ Households with a child born between September 2002 and August 2004 were identified from resident registries and recruited via random sampling (with oversampling of low-income households in anticipation of lower participation). From 2012-14 (T1), trained interviewers visited participant households every two years to administer questionnaires. Participating young people were approximately age 9-10 (T1, 2012-14), 11-12 (T2, 2014-16), 13-14 (T3, 2016-18), and 15-16 (T4, 2018-20) years old. 3,171 child-parent pairs entered the study at T1; 3,007 participated at T2 (94·8%), 2,667 at T3 (84·1%), and 2,616 at T4 (82·5%). Caregivers and young people gave informed written consent/assent at each wave. All procedures are approved by the Ethics Committees of the Tokyo Metropolitan Institute of Medical Science (no.: 12-35), the University of Tokyo (10057), and SOKENDAI (the Graduate University for Advanced Studies; 2012002).

REACH is an accelerated cohort study of adolescent mental health based in two inner-London boroughs, Lambeth and Southwark (www.thereachstudy.com).^14^ Three representative cohorts of young people – initially aged 11-12 (Cohort 1), 12-13 (Cohort 2), or 13-14 (Cohort 3) years – were recruited from 12 mainstream secondary schools (T1, 2016-17) and followed-up one (T2, 2017-18) and two (T3, 2018-19) years later. At each wave, young people provided informed written assent before completing computerised questionnaires. 4,353 participated at T1; 4,005 (92%) participated at T2 and/or T3. All procedures are approved by the Psychiatry, Nursing and Midwifery Research Ethics Subcommittee, King’s College London (15/16-2320).

We used the three waves of data from each study that maximise comparability of calendar years and ages, i.e., TTC: T2-T4; REACH T1-T3.

### Choice of primary measure

The Short Mood and Feelings Questionnaire (SMFQ) is a free, widely used, unidimensional 13-item self-report measure of depressive symptoms in the past two weeks.^16^ Example items include: ‘I cried a lot’ and ‘I felt miserable or unhappy’. Each item has three response options: true (0), sometimes true (1), not true (2), from which a total score (0-26) is calculated. Higher scores indicate greater depressive symptoms. The SMFQ was originally developed in English and validated against diagnoses of depression in US paediatric samples.^16^ The Japanese version was developed for TTC using back-translation. (To aid readers’ interpretation of effect size, longitudinal within-person analyses of REACH data suggest SMFQ scores increase by around two points on average in those who become a victim of bullying (*unpublished*).)

### Gender

In TTC, gender/sex (boy, girl) was obtained via resident register at recruitment. In REACH, gender/sex (boy, girl) was self-reported.

### Data analysis

First, we examined distributions and missingness on included variables, and produced summary statistics overall and by cohort, gender, and gender-within-cohort, at each wave. Second, we fit a series of ordinal-indicator multigroup and longitudinal confirmatory factor analysis (CFA) models to examine configural, metric (loadings), and scalar (thresholds) invariance across cohorts/places, genders, and ages/waves (Figures S1 and S2).^17^ To assess model fit, we considered fit indices in combination rather than in isolation, gave weight to indices that are not sensitive to large sample size, and used proposed cut-offs as a guide (i.e., RMSEA≤0·08, CFI,TLI>0·90, and SRMR<0·08 as acceptable fit; RMSEA<0·06, CFI,TLI≥0·95 as good fit).^17,18^ With the addition of each set of equality constraints, we considered a change in RMSEA (ΔRMSEA) <0·015, ΔCFI ≤0·010, and ΔSRMR ≤0·030 as support for invariance.^19^ This is consistent with the approach used by others with similar measures, ordinal indicators, large samples, and cross-cohort designs.^9,20^ As the 13 SMFQ items have 3-level ordinal response options, we used Weighted Least Squares Mean and Variance-adjusted (WLSMV) estimation (polychoric correlations) to estimate covariance matrices.

Third, we used latent growth curve models (LCMs) to (a) determine the best-fitting functional form (i.e., intercept-only vs linear growth), and (b) estimate mean trajectories of SMFQ scores from age 11 to age 16 years in boys and girls in each cohort. For each growth parameter (i.e., intercept and linear slope), we added a gender*cohort interaction to formally test our hypothesis that gender inequalities are larger among young people in London than in Tokyo. We used full information robust maximum likelihood to accommodate missing data and the non-normal distribution of SMFQ scores. In sensitivity analyses, we (a) repeated the LCM with square root transformed SMFQ scores, because kurtosis in the raw score distribution was high and toward or above generally accepted levels, and (b) re-ran the LCM with the REACH cohorts treated as three distinct groups (given the accelerated design). Analyses were conducted in Stata 17 and Mplus 8.

### Youth involvement

To co-write this paper with young people (see author list), we (1) held several meetings and workshops with five young people in London (age 18-19) and five in Tokyo (age 19-20) to discuss findings and explore possible interpretations, explanations, and implications, (2) integrate their perspectives throughout, and (3) present their ideas in the Supplement (S6).

## RESULTS

### Descriptive information

In total, 7,100 young people (TTC, 2,813; REACH, 4,287) were included in this analysis. Half were boys (*n*=3,587, 50·5%), half girls (*n*=3,513, 49·5%). In TTC, reasons for being excluded from analyses (*n*=358 of 3,171 who took part at T1) were: lost to follow-up after the first wave (*n*=164) or did not complete the SMFQ (*n*=194). In REACH, reasons for being excluded from analyses (*n*=658 of 4,945 invited to take part at T1) were: opted-out by parent/carer (*n*=160): child declined to participate (*n*=20); persistent absence from school (*n*=80); or did not complete the SMFQ (*n*=398). The included samples remain highly representative of the target populations based on socio-demographic indicators.^13,14^ The REACH sample comprises high proportions from ethnically minoritised groups (*n*=3613, 84%), over a quarter eligible for free school meals (an indicator of low household income; *n*=1,183, 28%), around half boys (n, 2097, 49%), and roughly even proportions from each of the three REACH cohorts (Cohort 1: *n*=1,580; Cohort 2: *n*=1,382; Cohort 3: *n*=1,325). The TTC sample comprises just over half boys (n, 1490, 53%), 9% (n=242) with a household income <4 million yen (an indicator of low household income; the tax-free threshold is just under 2 million yen per person), and most had two Japanese parents (n, 2616, 97%). Summary statistics for SMFQ items and scores at age 11–12, by cohort and gender, are presented (Table 1, Figure 1).

**Table 1.** Summary statistics for the thirteen SMFQ items, by cohort and gender, at age 11-12 years.

|  |  | TTC |  |  |  | REACH |  |  |  |
| --- | --- | --- | --- | --- | --- | --- | --- | --- | --- |
|  |  | BOYS |  | GIRLS |  | BOYS |  | GIRLS |  |
|  |  | n | % | n | % | n | % | n | % |
| SMFQ Item 1 | not true | 736 | 55.3 | 629 | 53.0 | 328 | 56.5 | 324 | 49.1 |
|  | sometimes true | 449 | 33.7 | 423 | 35.7 | 184 | 31.7 | 245 | 37.1 |
|  | always true | 146 | 11.0 | 134 | 11.3 | 69 | 11.9 | 91 | 13.8 |
| SMFQ Item 2 | not true | 1,066 | 80.2 | 1,000 | 84.3 | 436 | 74.9 | 471 | 71.6 |
|  | sometimes true | 215 | 16.2 | 157 | 13.2 | 100 | 17.2 | 156 | 23.7 |
|  | always true | 48 | 3.6 | 29 | 2.5 | 46 | 7.9 | 31 | 4.7 |
| SMFQ Item 3 | not true | 856 | 64.4 | 782 | 66.1 | 297 | 51.2 | 349 | 52.6 |
|  | sometimes true | 362 | 27.2 | 314 | 26.5 | 181 | 31.2 | 217 | 32.7 |
|  | always true | 112 | 8.4 | 87 | 7.4 | 102 | 17.6 | 97 | 14.6 |
| SMFQ Item 4 | not true | 900 | 67.6 | 881 | 74.3 | 356 | 61.5 | 447 | 67.8 |
|  | sometimes true | 333 | 25.0 | 235 | 19.8 | 157 | 27.1 | 149 | 22.6 |
|  | always true | 98 | 7.4 | 70 | 5.9 | 66 | 11.4 | 63 | 9.6 |
| SMFQ Item 5 | not true | 1,083 | 81.4 | 984 | 83.3 | 444 | 76.8 | 477 | 72.8 |
|  | sometimes true | 193 | 14.5 | 156 | 13.2 | 76 | 13.2 | 106 | 16.2 |
|  | always true | 54 | 4.1 | 42 | 3.6 | 58 | 10.0 | 72 | 11.0 |
| SMFQ Item 6 | not true | 1,186 | 89.5 | 975 | 82.4 | 476 | 83.8 | 456 | 69.1 |
|  | sometimes true | 109 | 8.2 | 148 | 12.5 | 51 | 9.0 | 141 | 21.4 |
|  | always true | 30 | 2.3 | 60 | 5.1 | 41 | 7.2 | 63 | 9.6 |
| SMFQ Item 7 | not true | 833 | 62.7 | 798 | 67.3 | 303 | 52.6 | 340 | 51.4 |
|  | sometimes true | 383 | 28.8 | 330 | 27.9 | 189 | 32.8 | 225 | 34.0 |
|  | always true | 112 | 8.4 | 57 | 4.8 | 84 | 14.6 | 96 | 14.5 |
| SMFQ Item 8 | not true | 1,079 | 81.2 | 866 | 73.1 | 463 | 80.8 | 503 | 76.8 |
|  | sometimes true | 190 | 14.3 | 231 | 19.5 | 65 | 11.3 | 96 | 14.7 |
|  | always true | 60 | 4.5 | 88 | 7.4 | 45 | 7.9 | 56 | 8.6 |
| SMFQ Item 9 | not true | 984 | 74.1 | 853 | 72.0 | 465 | 80.9 | 536 | 81.6 |
|  | sometimes true | 289 | 21.8 | 270 | 22.8 | 75 | 13.0 | 80 | 12.2 |
|  | always true | 55 | 4.1 | 61 | 5.2 | 35 | 6.1 | 41 | 6.2 |
| SMFQ Item 10 | not true | 1,153 | 86.8 | 962 | 81.3 | 440 | 76.7 | 459 | 69.8 |
|  | sometimes true | 132 | 9.9 | 163 | 13.8 | 91 | 15.9 | 132 | 20.1 |
|  | always true | 44 | 3.3 | 59 | 5.0 | 43 | 7.5 | 67 | 10.2 |
| SMFQ Item 11 | not true | 1,092 | 82.4 | 956 | 80.8 | 474 | 82.6 | 500 | 75.9 |
|  | sometimes true | 176 | 13.3 | 161 | 13.6 | 56 | 9.8 | 102 | 15.5 |
|  | always true | 58 | 4.4 | 66 | 5.6 | 44 | 7.7 | 57 | 8.7 |
| SMFQ Item 12 | not true | 1,063 | 80.1 | 927 | 78.5 | 426 | 74.1 | 449 | 68.3 |
|  | sometimes true | 211 | 15.9 | 187 | 15.8 | 98 | 17.0 | 138 | 21.0 |
|  | always true | 53 | 4.0 | 67 | 5.7 | 51 | 8.9 | 70 | 10.7 |
| SMFQ Item 13 | not true | 1,096 | 82.53 | 962 | 81.25 | 450 | 78.8 | 512 | 78.9 |
|  | sometimes true | 194 | 14.61 | 187 | 15.79 | 84 | 14.7 | 93 | 14.3 |
|  | always true | 38 | 2.86 | 35 | 2.96 | 37 | 6.5 | 45 | 6.9 |

### Measurement invariance

The unidimensional factor structure was well supported in both cohorts and genders, and at all waves/ages. For example, in the cross-cohort multigroup CFA at age 11-12 years, the configural model fit well (CFI, 0·970; TLI, 0·965; SRMR, 0·048; RMSEA, 0·064; Table 2) and all item loadings were high (0·6 to 0·9) in both cohorts (Table S3). Overall, loading and threshold (i.e., scalar) invariance across cohorts and genders was well supported (Table 2). For instance, constraining item loadings and thresholds to be equal across cohorts at age 11-12 led to change in fit that was within the acceptable range across all fit indices (e.g., SRMR changed from 0·048 [configural], to 0·050 [metric], to 0·051 [scalar]), and the overall fit of the scalar model was good (CFI 0·964, RMSEA 0·065). Support for scalar invariance was generally strong across all fit indices in each CFA, with two exceptions: (a) in cross-cohort invariance tests at age 15-16, ΔCFI and ΔSRMR were comfortably within the acceptable range, whereas ΔRMSEA for the metric model was at the upper limit of what is generally considered acceptable (ΔRMSEA: 0·016); and (b) in models examining invariance by gender within REACH at age 15-16, the overall fit of the scalar model was good according to most indices (i.e., CFI and TLI 0·969, SRMR 0·066) but at the upper limit based on the RMSEA (0·090, 90% CI: 0·083, 0·097)). Longitudinal invariance of loadings and thresholds over time within each cohort was also well-supported (Table 3; Mplus syntax provided in S4). For example, in TTC, change in fit was comfortably within the acceptable range when constraining loadings and thresholds to be equal over time (e.g., ΔCFI 0·004; ΔRMSEA 0·002).

**Table 2.**
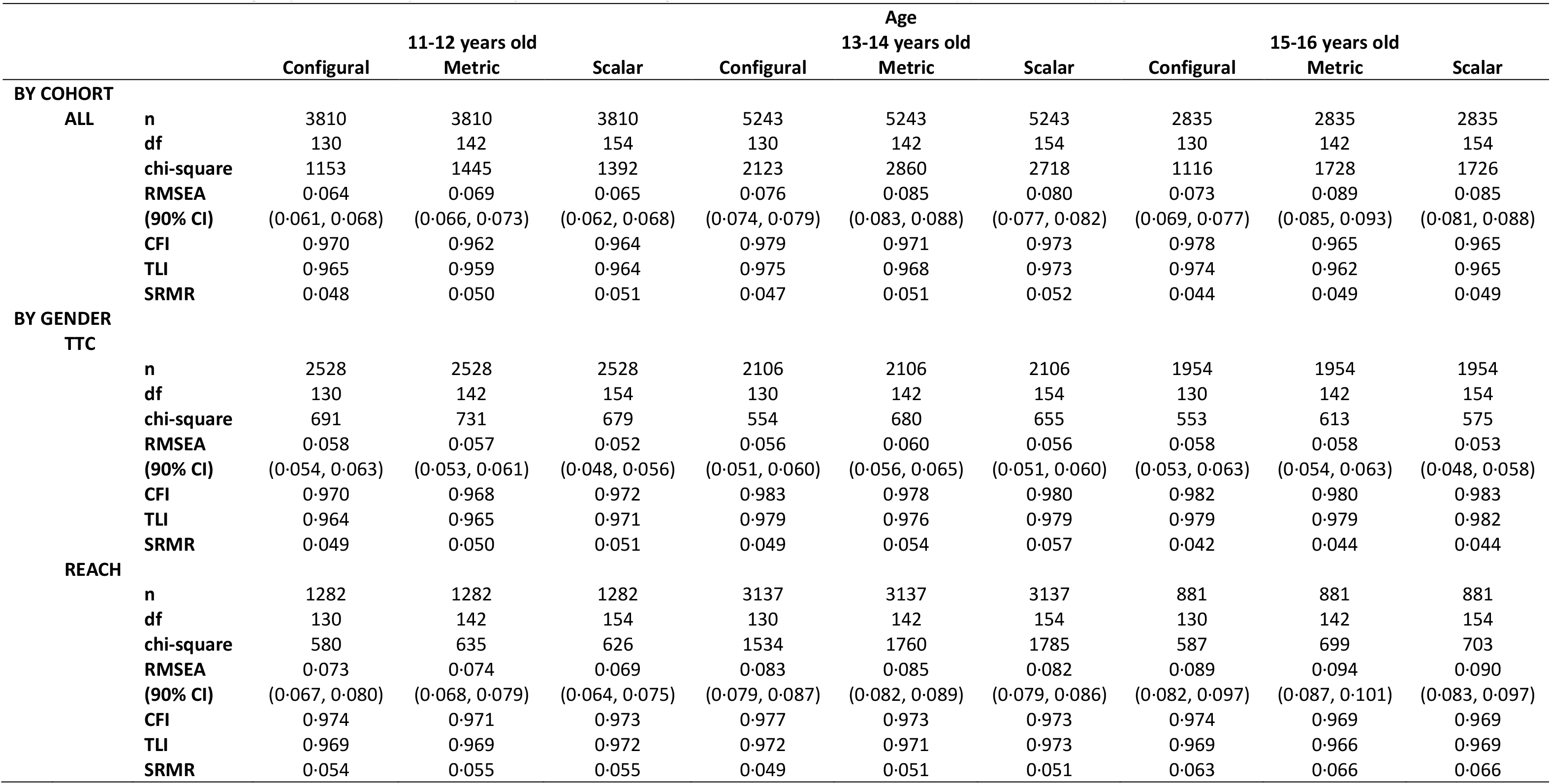
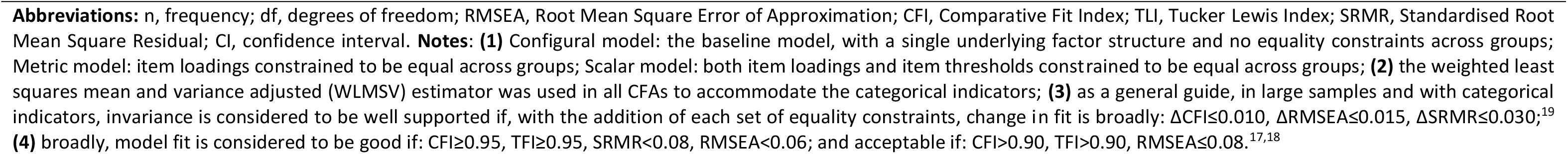
Fit indices for multigroup confirmatory factor analysis models testing measurement invariance across (a) cohorts, and (b) genders.

|  |  | 11-12 years old |  |  | Age<br>13-14 years old |  |  | 15-16 years old |  |  |
| --- | --- | --- | --- | --- | --- | --- | --- | --- | --- | --- |
|  |  | Configural | Metric | Scalar | Configural | Metric | Scalar | Configural | Metric | Scalar |
| **BY COHORT** |  |  |  |  |  |  |  |  |  |  |
| **ALL** | **n** | 3810 | 3810 | 3810 | 5243 | 5243 | 5243 | 2835 | 2835 | 2835 |
|  | **df** | 130 | 142 | 154 | 130 | 142 | 154 | 130 | 142 | 154 |
|  | **chi-square** | 1153 | 1445 | 1392 | 2123 | 2860 | 2718 | 1116 | 1728 | 1726 |
|  | **RMSEA** | 0.064 | 0.069 | 0.065 | 0.076 | 0.085 | 0.080 | 0.073 | 0.089 | 0.085 |
|  | **(90% CI)** | (0.061, 0.068) | (0.066, 0.073) | (0.062, 0.068) | (0.074, 0.079) | (0.083, 0.088) | (0.077, 0.082) | (0.069, 0.077) | (0.085, 0.093) | (0.081, 0.088) |
|  | **CFI** | 0.970 | 0.962 | 0.964 | 0.979 | 0.971 | 0.973 | 0.978 | 0.965 | 0.965 |
|  | **TLI** | 0.965 | 0.959 | 0.964 | 0.975 | 0.968 | 0.973 | 0.974 | 0.962 | 0.965 |
|  | **SRMR** | 0.048 | 0.050 | 0.051 | 0.047 | 0.051 | 0.052 | 0.044 | 0.049 | 0.049 |
| **BY GENDER** |  |  |  |  |  |  |  |  |  |  |
| **TTC** | **n** | 2528 | 2528 | 2528 | 2106 | 2106 | 2106 | 1954 | 1954 | 1954 |
|  | **df** | 130 | 142 | 154 | 130 | 142 | 154 | 130 | 142 | 154 |
|  | **chi-square** | 691 | 731 | 679 | 554 | 680 | 655 | 553 | 613 | 575 |
|  | **RMSEA** | 0.058 | 0.057 | 0.052 | 0.056 | 0.060 | 0.056 | 0.058 | 0.058 | 0.053 |
|  | **(90% CI)** | (0.054, 0.063) | (0.053, 0.061) | (0.048, 0.056) | (0.051, 0.060) | (0.056, 0.065) | (0.051, 0.060) | (0.053, 0.063) | (0.054, 0.063) | (0.048, 0.058) |
|  | **CFI** | 0.970 | 0.968 | 0.972 | 0.983 | 0.978 | 0.980 | 0.982 | 0.980 | 0.983 |
|  | **TLI** | 0.964 | 0.965 | 0.971 | 0.979 | 0.976 | 0.979 | 0.979 | 0.979 | 0.982 |
|  | **SRMR** | 0.049 | 0.050 | 0.051 | 0.049 | 0.054 | 0.057 | 0.042 | 0.044 | 0.044 |
| **REACH** | **n** | 1282 | 1282 | 1282 | 3137 | 3137 | 3137 | 881 | 881 | 881 |
|  | **df** | 130 | 142 | 154 | 130 | 142 | 154 | 130 | 142 | 154 |
|  | **chi-square** | 580 | 635 | 626 | 1534 | 1760 | 1785 | 587 | 699 | 703 |
|  | **RMSEA** | 0.073 | 0.074 | 0.069 | 0.083 | 0.085 | 0.082 | 0.089 | 0.094 | 0.090 |
|  | **(90% CI)** | (0.067, 0.080) | (0.068, 0.079) | (0.064, 0.075) | (0.079, 0.087) | (0.082, 0.089) | (0.079, 0.086) | (0.082, 0.097) | (0.087, 0.101) | (0.083, 0.097) |
|  | **CFI** | 0.974 | 0.971 | 0.973 | 0.977 | 0.973 | 0.973 | 0.974 | 0.969 | 0.969 |
|  | **TLI** | 0.969 | 0.969 | 0.972 | 0.972 | 0.971 | 0.973 | 0.969 | 0.966 | 0.969 |
|  | **SRMR** | 0.054 | 0.055 | 0.055 | 0.049 | 0.051 | 0.051 | 0.063 | 0.066 | 0.066 |

**Abbreviations:** n, frequency; df, degrees of freedom; RMSEA, Root Mean Square Error of Approximation; CFI, Comparative Fit Index; TLI, Tucker Lewis Index; SRMR, Standardised Root Mean Square Residual; CI, confidence interval. **Notes:** **(1)** Configural model: the baseline model, with a single underlying factor structure and no equality constraints across groups; Metric model: item loadings constrained to be equal across groups; Scalar model: both item loadings and item thresholds constrained to be equal across groups; **(2)** the weighted least squares mean and variance adjusted (WLSMV) estimator was used in all CFAs to accommodate the categorical indicators; **(3)** as a general guide, in large samples and with categorical indicators, invariance is considered to be well supported if, with the addition of each set of equality constraints, change in fit is broadly: $\Delta CFI \leq 0.010$ , $\Delta RMSEA \leq 0.015$ , $\Delta SRMR \leq 0.030$ ; <sup>19</sup> **(4)** broadly, model fit is considered to be good if: $CFI \geq 0.95$ , $TLI \geq 0.95$ , $SRMR < 0.08$ , $RMSEA < 0.06$ ; and acceptable if: $CFI > 0.90$ , $TLI > 0.90$ , $RMSEA \leq 0.08$ .<sup>17,18</sup>

**Table 3.**
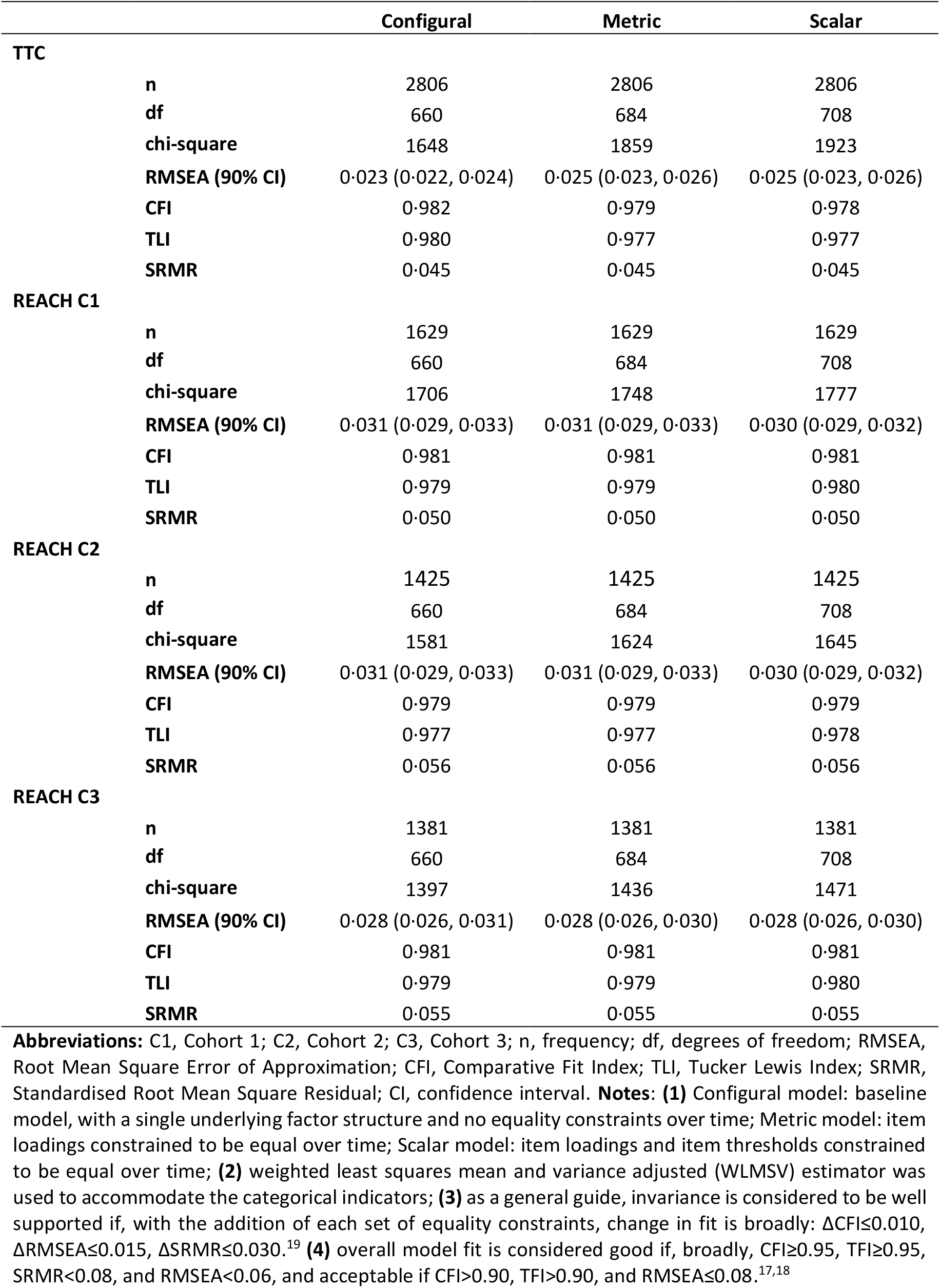
Fit indices for longitudinal confirmatory factor analysis models testing measurement invariance over time within each cohort.

|  | Configural | Metric | Scalar |
| --- | --- | --- | --- |
| **TTC** |  |  |  |
| n | 2806 | 2806 | 2806 |
| df | 660 | 684 | 708 |
| chi-square | 1648 | 1859 | 1923 |
| RMSEA (90% CI) | 0.023 (0.022, 0.024) | 0.025 (0.023, 0.026) | 0.025 (0.023, 0.026) |
| CFI | 0.982 | 0.979 | 0.978 |
| TLI | 0.980 | 0.977 | 0.977 |
| SRMR | 0.045 | 0.045 | 0.045 |
| **REACH C1** |  |  |  |
| n | 1629 | 1629 | 1629 |
| df | 660 | 684 | 708 |
| chi-square | 1706 | 1748 | 1777 |
| RMSEA (90% CI) | 0.031 (0.029, 0.033) | 0.031 (0.029, 0.033) | 0.030 (0.029, 0.032) |
| CFI | 0.981 | 0.981 | 0.981 |
| TLI | 0.979 | 0.979 | 0.980 |
| SRMR | 0.050 | 0.050 | 0.050 |
| **REACH C2** |  |  |  |
| n | 1425 | 1425 | 1425 |
| df | 660 | 684 | 708 |
| chi-square | 1581 | 1624 | 1645 |
| RMSEA (90% CI) | 0.031 (0.029, 0.033) | 0.031 (0.029, 0.033) | 0.030 (0.029, 0.032) |
| CFI | 0.979 | 0.979 | 0.979 |
| TLI | 0.977 | 0.977 | 0.978 |
| SRMR | 0.056 | 0.056 | 0.056 |
| **REACH C3** |  |  |  |
| n | 1381 | 1381 | 1381 |
| df | 660 | 684 | 708 |
| chi-square | 1397 | 1436 | 1471 |
| RMSEA (90% CI) | 0.028 (0.026, 0.031) | 0.028 (0.026, 0.030) | 0.028 (0.026, 0.030) |
| CFI | 0.981 | 0.981 | 0.981 |
| TLI | 0.979 | 0.979 | 0.980 |
| SRMR | 0.055 | 0.055 | 0.055 |

**Abbreviations:** C1, Cohort 1; C2, Cohort 2; C3, Cohort 3; n, frequency; df, degrees of freedom; RMSEA, Root Mean Square Error of Approximation; CFI, Comparative Fit Index; TLI, Tucker Lewis Index; SRMR, Standardised Root Mean Square Residual; CI, confidence interval. **Notes:** **(1)** Configural model: baseline model, with a single underlying factor structure and no equality constraints over time; Metric model: item loadings constrained to be equal over time; Scalar model: item loadings and item thresholds constrained to be equal over time; **(2)** weighted least squares mean and variance adjusted (WLSMV) estimator was used to accommodate the categorical indicators; **(3)** as a general guide, invariance is considered to be well supported if, with the addition of each set of equality constraints, change in fit is broadly: $\Delta CFI \leq 0.010$ , $\Delta RMSEA \leq 0.015$ , $\Delta SRMR \leq 0.030$ .<sup>19</sup> **(4)** overall model fit is considered good if, broadly, $CFI \geq 0.95$ , $TFI \geq 0.95$ , $SRMR < 0.08$ , and $RMSEA < 0.06$ , and acceptable if $CFI > 0.90$ , $TFI > 0.90$ , and $RMSEA \leq 0.08$ .<sup>17,18</sup>

### Depressive symptom trajectories

#### Overall, by cohort

At each age/wave, there was strong evidence of lower mean depressive symptoms among young people in Tokyo compared with young people in London (Figure S5). The estimated average starting level (i.e., mean intercept) was 1·1 points lower (95% CI: -1·4, -0·8), and average rate of change per year (i.e., mean slope) was 0·7 (-0·8, -0·6) points lower among young people in Tokyo (Figure S5).

#### By cohort and gender

Among young people in London, there was strong evidence of a modest difference in mean depressive symptoms between boys and girls at age 11-12 years (difference in mean intercepts: 0·8 [0·3, 1·2]) (Figure 2). Among young people in Tokyo, there was no evidence of a difference between boys and girls at age 11-12 years (0·19 [-0·15, 0·53]) (gender-by-cohort interaction for the intercept: *p*=0·069); however, a difference emerged between ages 11 and 14 years, due to decreases with age among boys and small increases on average among girls (mean difference in slopes: 0·52 [0·40, 0·65]). In both cohorts, the disparity between boys and girls widened year-on-year, but there was strong evidence of a gender-by-cohort interaction in mean rate of change (*p*<0·001). In particular, the average rate of increase in depressive symptoms per year was around four times greater among girls in London (1·1 [0·9, 1·3]) compared with girls in Tokyo (0·3 [0·2, 0·4]), and by age 16 years, the difference between boys and girls was around twice as large among young people in London than in Tokyo.

#### Sensitivity analyses

Using transformed SMFQ scores and separating the three REACH cohorts made no substantive difference to our findings (Figures S7 and S8).

## DISCUSSION

### Key findings

We found strong evidence that gender inequalities in depressive symptoms are larger, and may emerge earlier, among young people in London than among young people in Tokyo. Notably, the mean rate of increase in depressive symptoms per year was around four times steeper among girls in London than among girls in Tokyo. We found little evidence to suggest these differences are due to incomparable measurement. Our work extends and strengthens a growing body of evidence on the context dependent nature of gender inequalities in emotional health in several important ways: robust consideration of measurement equivalence; the longitudinal design spanning the age at which gender inequalities in emotional health typically emerge; large representative samples of young people in places that, at nation-level, rank far apart on societal gender equity metrics; and the involvement of young people in writing this paper.

### Limitations

Nonetheless, several limitations should be noted. First, gender is not a binary construct, however the available cross-cohort data limited our investigation to comparisons of boys and girls. We use the term gender, rather than sex, because our hypotheses are grounded in social (not sex-linked biological) causes of gender inequalities in mental health. Second, being limited to three waves meant we were only able to model linear trajectories. With additional waves of data, it will become possible to model non-linear growth, which may provide a better fit in TTC. Third, while measurement invariance tests can identify individual items or groups of items that function differently across groups, it cannot detect systematic under-reporting on *each and every* item. As highlighted by young people in our team (see S6), this is an important caveat. The extent to which this potential issue contributes to the differences we report, and in which direction(s), is unclear; cross-cohort qualitative work may be a useful next step.

### Findings in context

Our findings are consistent with a growing body of evidence on the context-dependent nature of gender inequalities in mental health.^5,7,9^ Using cross-sectional data on over 500,000 adolescents aged 15 years from 73 countries, Campbell and colleagues (2021) found substantial variation in the size of gender inequalities in mental distress and mental wellbeing across countries.^9^ Consistent with our findings, they also found that gender inequalities in mental distress tend to be *larger* in countries that perform *better* on formal indices of societal gender equity. In their analyses, the UK (currently ranked 15^th^ in the GGGI^15^) had one of the largest gender distress gaps, whereas Japan (ranked 125^th^) had one of the smallest.

Collectively, the evidence suggests there are social and structural drivers underpinning elevated rates of emotional distress among women and girls. But how might we explain the paradoxical finding that ostensibly more gender-equal societies have greater gender inequalities in depression? In relation to adults, leading theories include ‘mismatch between reality and expectation’ and ‘role overload’. While social expectations for women to succeed in education and employment have greatly increased, traditional attitudes placing domestic work and childcare responsibilities predominantly on women remain firmly entrenched.^21^ As noted by our Young Researchers, indices like the GGGI are limited as they only capture certain aspects of gender inequity; they do not paint the full picture. The GGGI assesses gender (in)equity in health, education, economic participation and political representation, but does not include things like unpaid work, gender-norm attitudes, and gender-based violence. Sexual harassment, assault, and intimate partner violence (IPV), are strongly associated with mental ill-health and are common in countries that perform well on the GGGI^15^, including among young people.^8^ Though comparable data is scarce, there are indeed indications that prevalence of and attitudes toward gender-based violence is better in Japan than in the UK/Great Britain (GB), e.g., lifetime prevalence of IPV is lower, as is the proportion of women who agree that IPV is ‘justified in certain circumstances’.^22^

While such factors are evidently relevant to older adolescents and young adults (e.g., 16-25-year-olds engaged in unpaid care are disproportionately female^23^ and, as discussed earlier, sexual violence explains part of the gender mental health gap in mid-to-late adolescence^8^), it is less clear how such factors could explain the emergence of gender inequalities in mental health by age 11. In part, this is because we lack data on relevant social experience (e.g., domestic work, gender-based violence, gendered parenting, etc.) among girls of this age; most of the research in this area has focussed on biological explanations, particularly those related to puberty.^1,2^ Evidence that sex hormones are responsible for the divergence of emotional health between boys and girls is inconsistent, correlational at best, and the mechanisms are unknown.^24^ Very few studies have investigated the possibility that the mechanism linking puberty to the divergence of emotional health between boys and girls in early adolescence could be *exogenous*, e.g., driven by the way society treats girls as they develop.^25^ Consequently, potentially modifiable targets to prevent gender inequalities in mental health from emerging at this age remain under-investigated, under-theorised, and unidentified.

Among our Young Researchers, there was a strong sense that young people in London may assume ‘adult’ roles and responsibilities at a younger age compared with young people in Tokyo who may typically be more ‘protected’ from such responsibilities during high school. In the UK, young girls and those in low income and financially insecure families do assume ‘adult’ responsibilities such as unpaid care at younger ages than do boys and those from more affluent backgrounds.^23^ Given the relative social disadvantage of young people in REACH compared with TTC, it is possible that (gendered) differences in young people’s roles and responsibilities in the home and the community contribute to disproportionately high depressive symptoms among girls in London. Considering Japan’s high gender equity in health and secondary education outcomes but low equity in economic and political participation,^15^ we speculate that in Tokyo the gender depression gap will widen *later*, around early adulthood, as young women who experience comparatively equal treatment throughout compulsory education are confronted with structural barriers in work, career advancement, and childcare. We will test this hypothesis when the cohort reaches adulthood.

In the UK, there is much concern about the possible impacts of social media use and academic pressure on the mental health of young people, particularly girls. We were unable to find comparable data for our two cities/countries, but discussions with our Young Researchers suggest social media use is ubiquitous – and perhaps similar in nature – in both places, and more broadly we note that gender inequalities in emotional health predate the invention of social media and smartphones.^1^ However, we cannot rule out that differences in use and impacts may contribute to our findings. A recent systematic review of qualitative evidence in the UK suggests that academic expectations and pressures are greater among girls than among boys.^26^ Our Young Researchers suggest academic pressure is universally high in Japan, potentially reducing the gender disparity in comparison, though again comparable data is scarce.

Finally, societal and structural differences may contribute to the overall higher levels of depressive symptoms among young people in London compared with Tokyo: violence and interpersonal crime is significantly lower in Tokyo;^27^ experience of racism is common among young people in London^28^ (and London is far more ethnically diverse than Tokyo); and the UK’s prolonged period of austerity – and related increases in child poverty and inequality and chronic underfunding of public services supporting young people – is not mirrored in Japan. These interlinked structural harms are strongly associated with mental distress among young people.^28,29^ Whether they also contribute to larger gender inequality in depressive symptoms and steeper increases in emotional distress among girls in London is unclear. We will explore the possibility of harmonising data on social conditions and experiences in TTC and REACH to investigate the likely confluence of factors that may explain our findings.

### Implications

In public debate, differences in emotional health between teenage boys and girls are often considered a fact of life, an inevitable part of teenage development. Our own and others’ work increasingly suggests this may not be case. If high and rising rates of emotional ill-health among girls and young women^30^ are largely driven by social and structural factors, then individual-focussed interventions (e.g., mindfulness, talking therapies, mental health first aid) – which currently dominate the UK’s response to the problem – are unlikely to be an effective solution.

### Conclusions

Our findings suggest gender inequalities in emotional health are context dependent and the steep increases in emotional distress experienced by many girls in the UK are not an inevitable part of development. There is an urgent need to understand the contexts and conditions that enable young girls to thrive.

## Data Availability

TTC: Data from TTC is archived in the Tokyo Metropolitan Institute of Medical Science. Collaboration in data analysis and publication will be welcome through specific research proposals sent to the research committee. The initial contact point for collaborations is Dr Atsushi Nishida.
REACH: We welcome requests from researchers wishing to access REACH data for specific research projects or collaborations. Our data access policy, which aims to make REACH data as accessible as possible while adhering to legal and ethical principles and protecting the privacy of schools and participants, can be found at www.thereachstudy.com/information-for-researchers.html. The application should be submitted to Professor Craig Morgan.

## CONTRIBUTORS

GK conceived, designed, and conducted the analyses, with input from DS, SY, CM & AN. GK and DS conducted the literature review and wrote the first and final drafts. KE, DS, GK, and CGA prepared the dataset for analysis. SU reviewed the analyses and provided expert statistical guidance. CGA, SD, DS, KL, KS, CM & GK collected and cleaned REACH data. Our young researchers (co-authors JK, AH, NT, KSCG, TR, and TTC’s Young Persons Advisory Group) were supported by and worked closely with MM, SY, KE, GK, EP, AN & DS. All authors contributed to interpretation of data, reviewed and critically revised the manuscript, and approved the final manuscript as submitted.

## DATA SHARING

TTC: Data from TTC is archived in the Tokyo Metropolitan Institute of Medical Science. Collaboration in data analysis and publication will be welcome through specific research proposals sent to the research committee. The initial contact point for collaborations is Dr Atsushi Nishida.

REACH: We welcome requests from researchers wishing to access REACH data for specific research projects or collaborations. Our data access policy, which aims to make REACH data as accessible as possible while adhering to legal and ethical principles and protecting the privacy of schools and participants, can be found at www.thereachstudy.com/information-for-researchers.html. The application should be submitted to Professor Craig Morgan.

## ACKNOWLEDGEMENTS

This work was supported by funding from the Invitation Program for Foreign Researchers at the Tokyo Metropolitan Institute of Medical Science. REACH is supported by the UK Economic and Social Research Council (ESRC) Centre for Society and Mental Health at King’s College London (ES/S012567/1); and the European Research Council (ERC) (REACH 648837). TTC is supported by funding from Grant-in-Aid for Scientific Research on Innovative Areas (23118002; Adolescent Mind & Self-Regulation) from the Ministry of Education, Culture, Sports, Science and Technology of Japan; JSPS KAKENHI (Grant Numbers JP16HY06395, 16H06398, 16H06399, 16K21720, 16K15566, 17H05931, JP21H05171, JP21H05173, JP21H05174 and JP22H05211); JST-Mirai Programme (Grant number JPMJMI21J3); and the International Research Center for Neurointelligence (WPI-IRCN) at the University of Tokyo Institutes for Advanced Study (UTIAS). We would like to thank all the young people, parents, teachers, and schools who generously contribute their time, information, and support to make REACH and TTC possible, and the students and staff who have contributed to data collection.

## Supplementary Material

**Figure S1.**
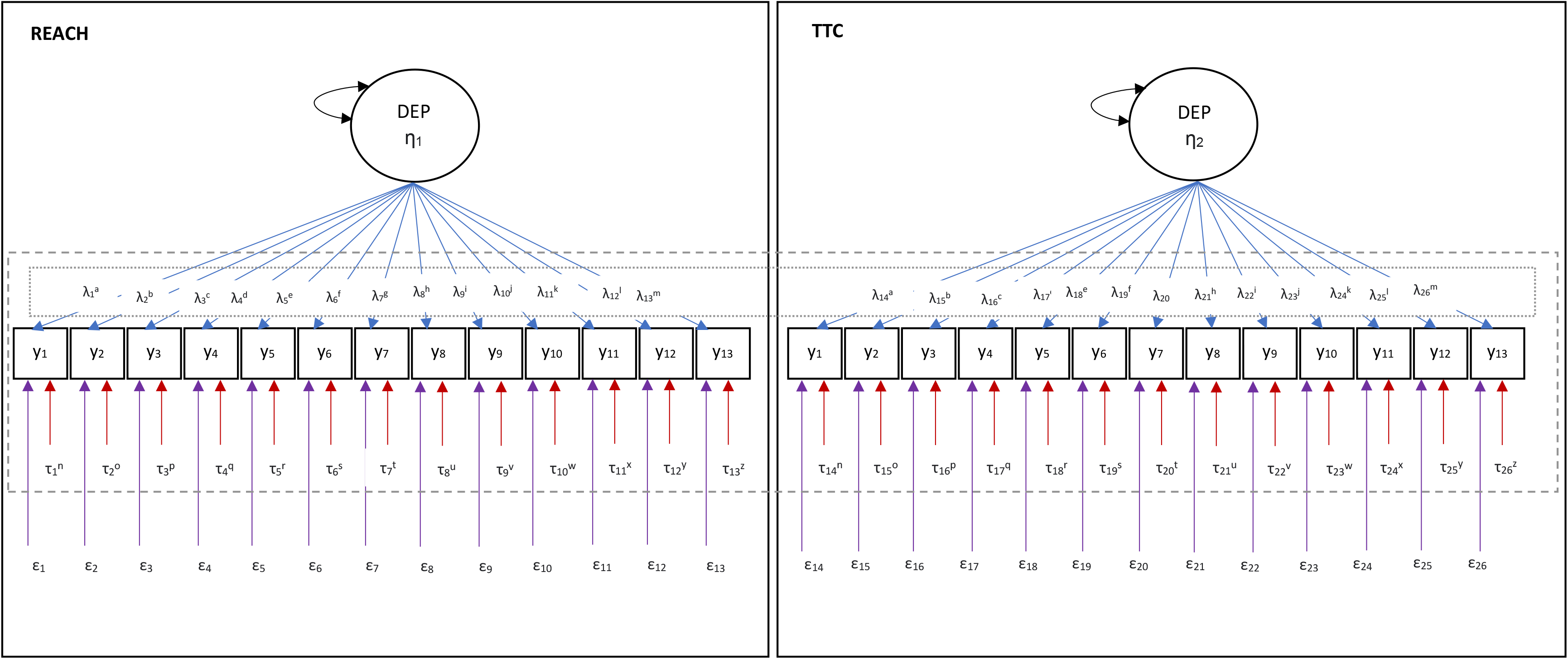
Path diagram for the ordinal-indicator multigroup confirmatory factor analysis model testing scalar invariance of the SMFQ across groups (i.e., cohorts, gender). Abbreviations: TTC, Tokyo Teen Cohort; REACH, Resilience, Ethnicity and AdolesCent Mental Health; DEP η, latent construct of depressive symptoms; y, item (indicator); λ, loading; τ, threshold; ε, residual. In the metric model, a-m loadings were constrained to be equal across groups. In the scalar model, a-n loadings and n-z thresholds were constrained to be equal across groups. The latent variable (DEP) mean was fixed to be one in Group 1 (here, REACH) and freely estimated in group 2 (here, TTC). Factor variances were freely estimated in both groups. The y1 factor loading was fixed at 1 in both groups (marker variable). Adapted from McElroy et al (2020).^20^

**Figure S2.**
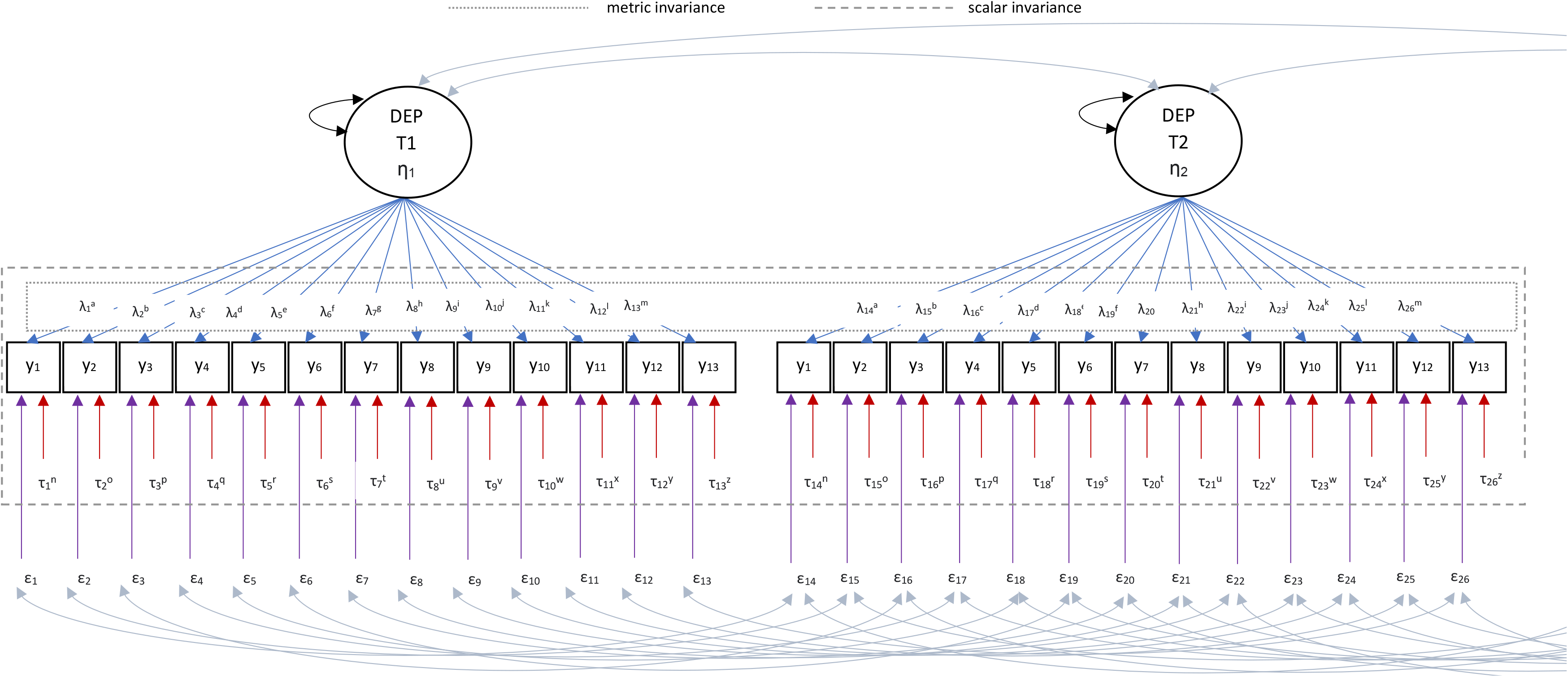
Path diagram for the longitudinal ordinal-indicator confirmatory factor analysis model testing scalar invariance of the SMFQ over time. (Note, figure truncated to two timepoints, but applied across all timepoints)

Abbreviations: T, timepoint/wave; DEP η, latent construct of depressive symptoms; y, item (indicator); λ, loading; τ, threshold; ε, residual. In the metric model, a-m loadings were constrained to be equal across timepoints. In the scalar model, a-n loadings and n-z thresholds were constrained to be equal across waves/timepoints. The latent variable (DEP) mean was fixed to be one at T1 and freely estimated at other timepoints. Factor variances were freely estimated at all timepoints. The y1 factor loading was fixed at 1 at all timepoints (marker variable). Adapted from McElroy et al (2020)20 and Liu et al (2017).^17^

**Table S3.**
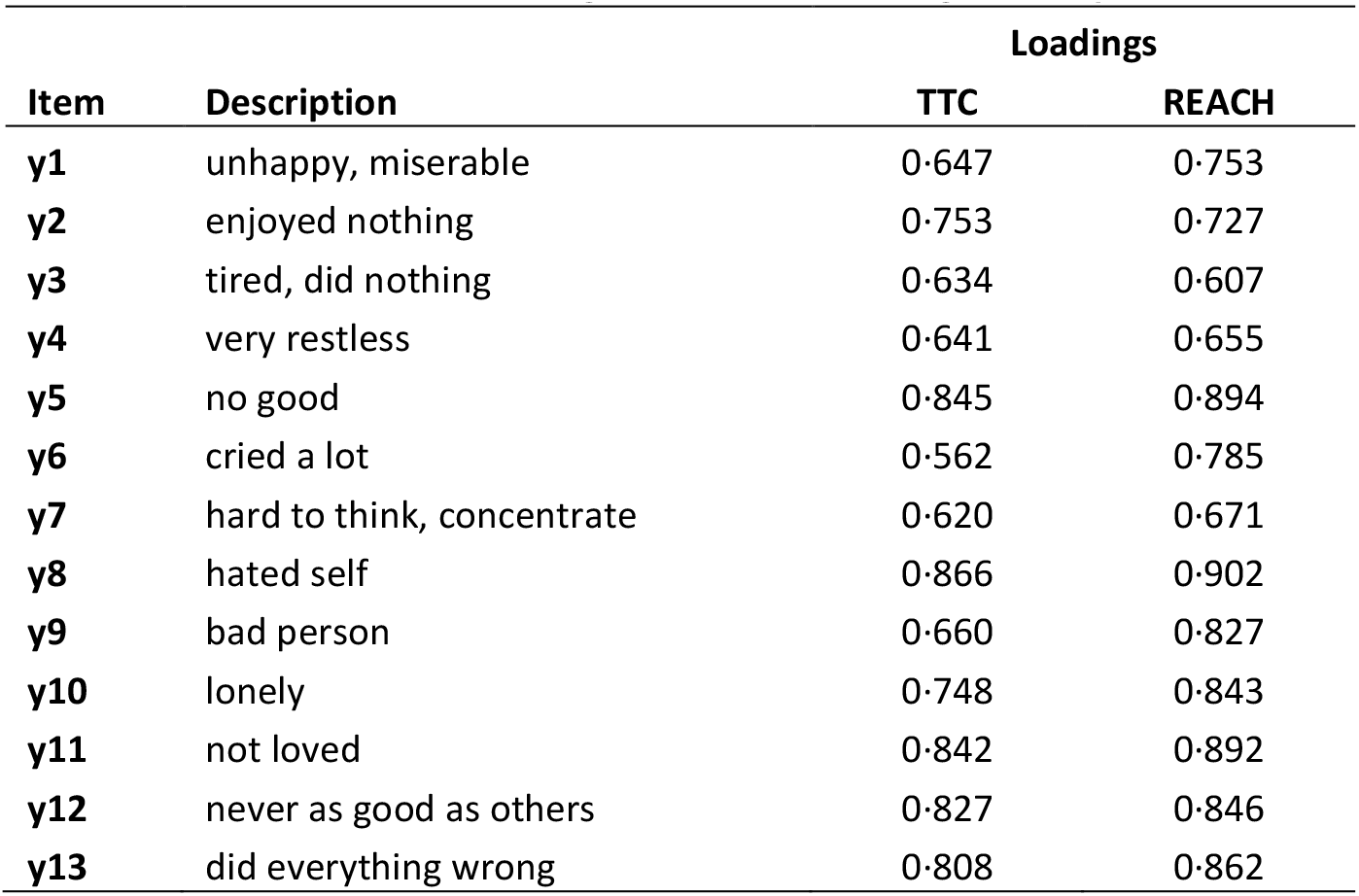
Estimated factor loadings in each cohort at age 11-12 years.

| Item | Description | Loadings |  |
| --- | --- | --- | --- |
|  |  | TTC | REACH |
| y1 | unhappy, miserable | 0.647 | 0.753 |
| y2 | enjoyed nothing | 0.753 | 0.727 |
| y3 | tired, did nothing | 0.634 | 0.607 |
| y4 | very restless | 0.641 | 0.655 |
| y5 | no good | 0.845 | 0.894 |
| y6 | cried a lot | 0.562 | 0.785 |
| y7 | hard to think, concentrate | 0.620 | 0.671 |
| y8 | hated self | 0.866 | 0.902 |
| y9 | bad person | 0.660 | 0.827 |
| y10 | lonely | 0.748 | 0.843 |
| y11 | not loved | 0.842 | 0.892 |
| y12 | never as good as others | 0.827 | 0.846 |
| y13 | did everything wrong | 0.808 | 0.862 |

**Figure S4.**
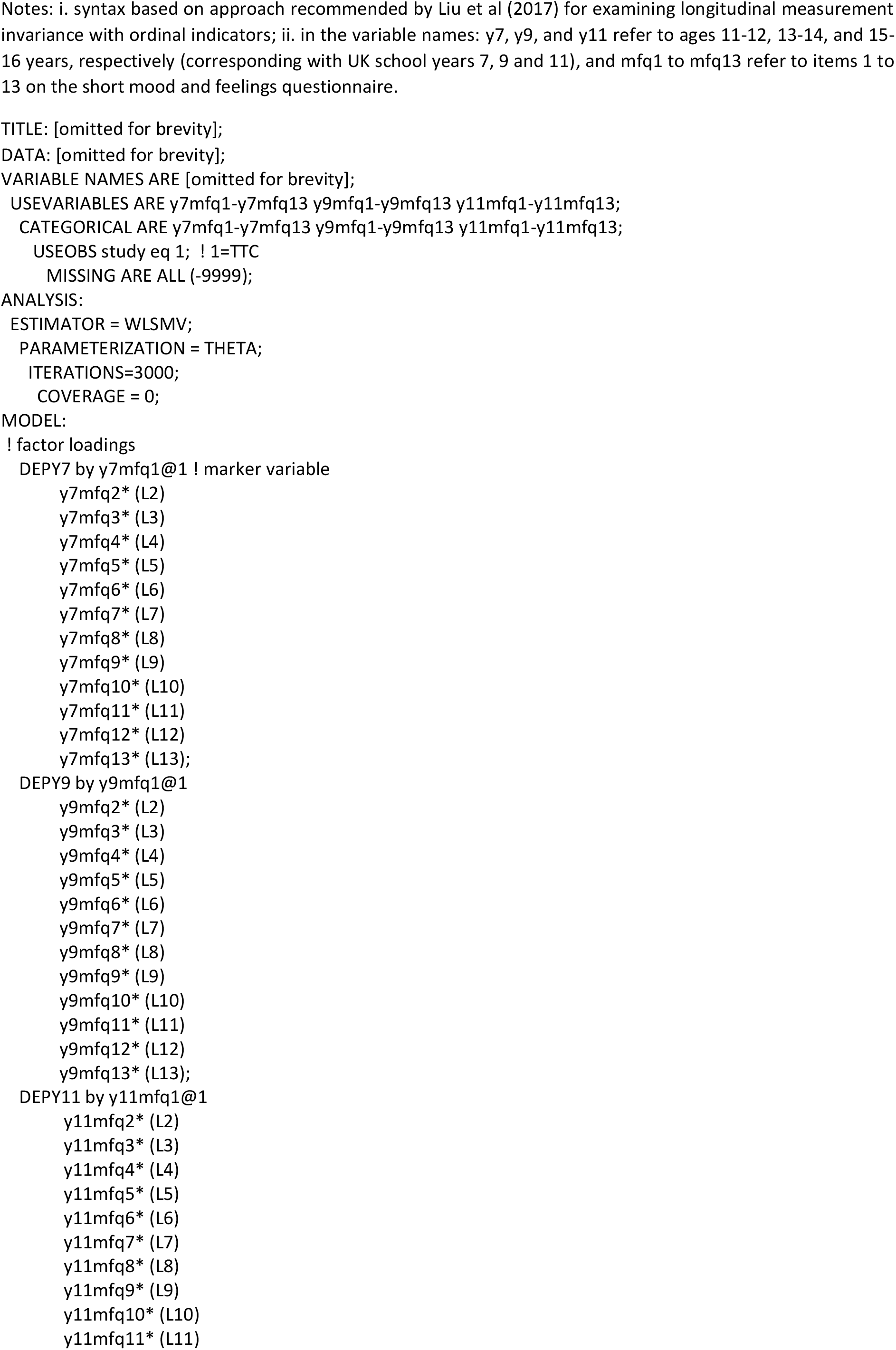
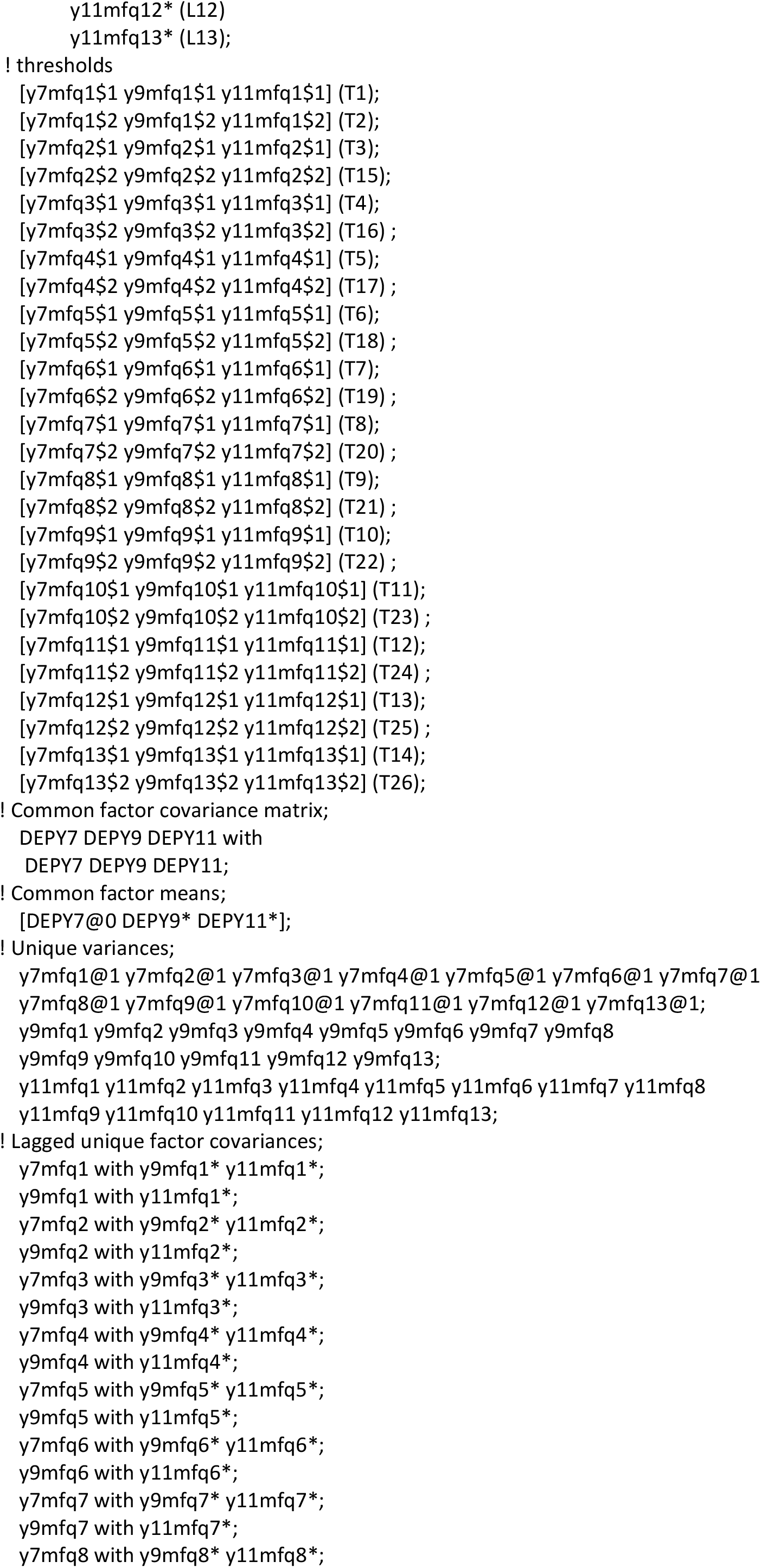
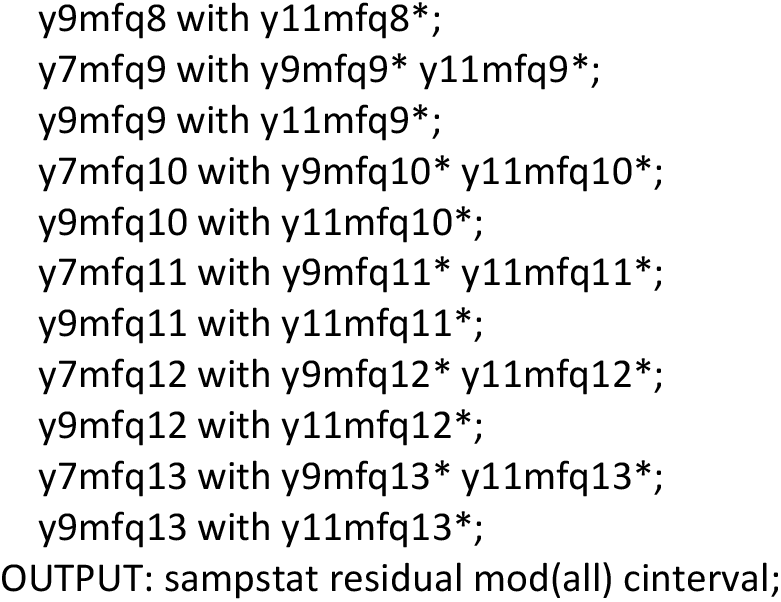
Mplus syntax for longitudinal confirmatory factor analysis in TTC: scalar model.

**Figure S5.**
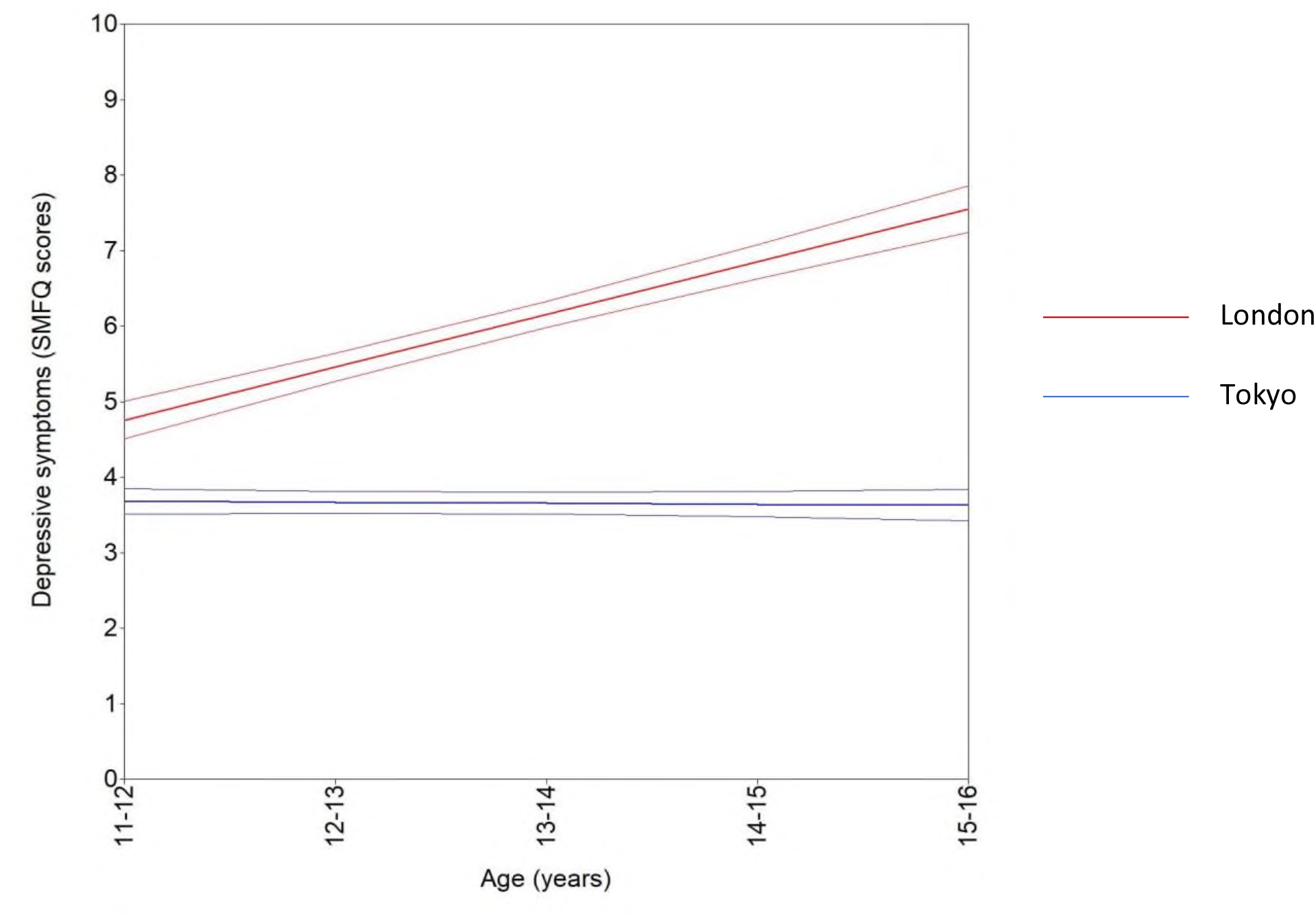
Estimated mean depressive symptoms trajectories (with 95% CI), by cohort (n, 7100).

**Figure S6.**
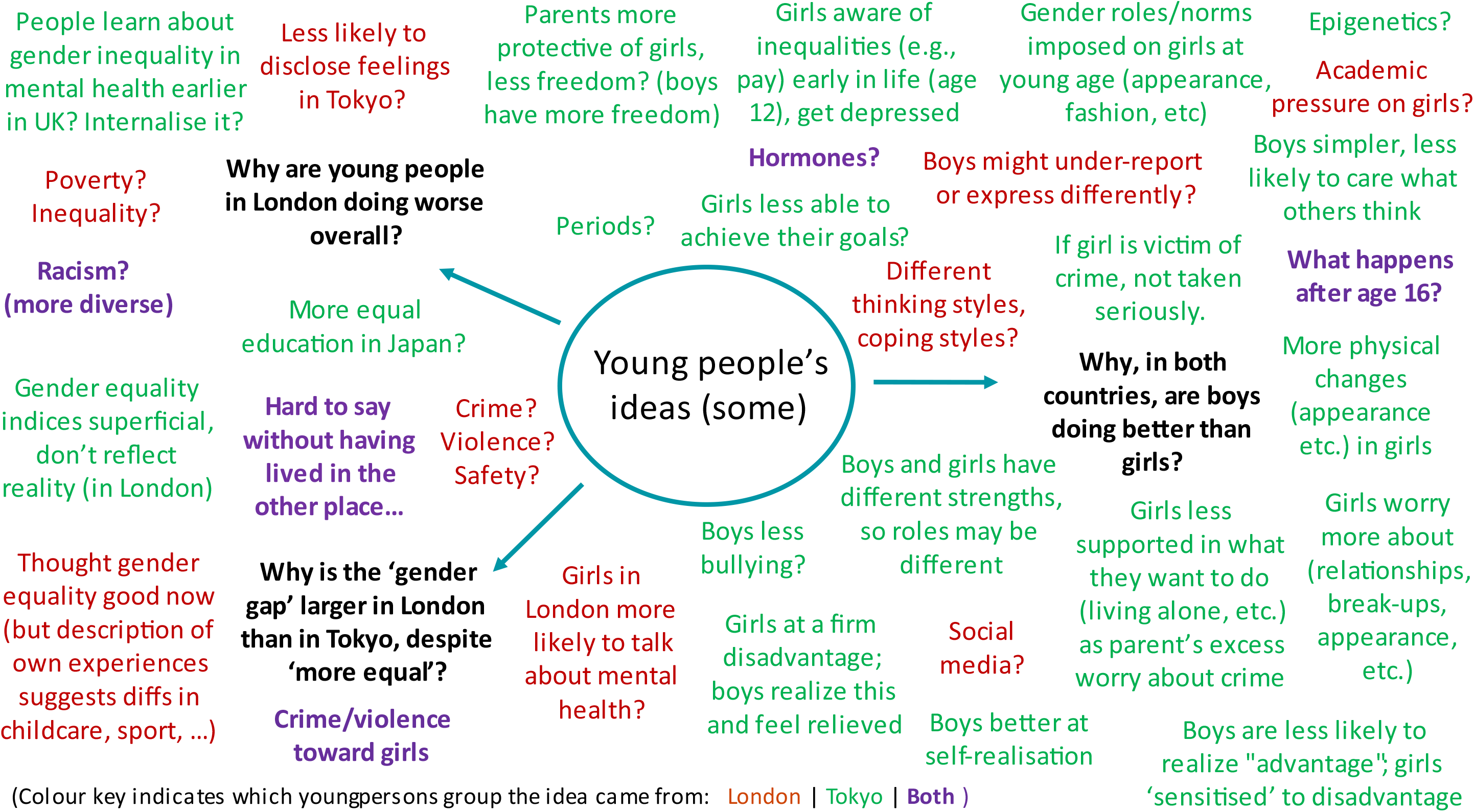
Mind map summarising ideas and reflections of young people in our team (see author list), presented in (a) English, and (b) Japanese. [Note, a Japanese version of the young persons mindmap is available on request from the authors. We were not able to upload it as part of this pre-print.

**Figure S7.**
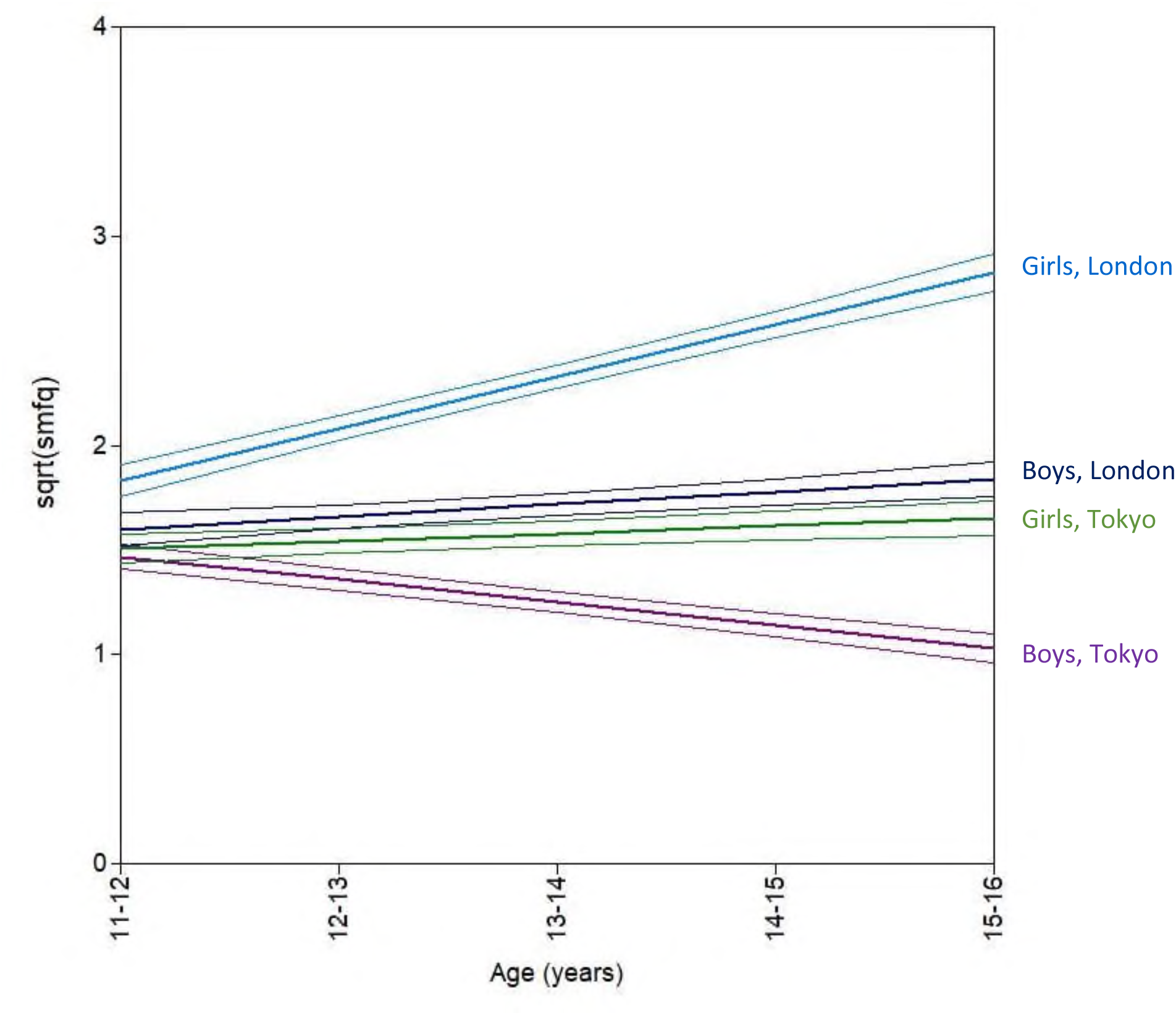
Latent growth curve model with square root transformed SMFQ scores as the dependent variable.

**Figure S8.**
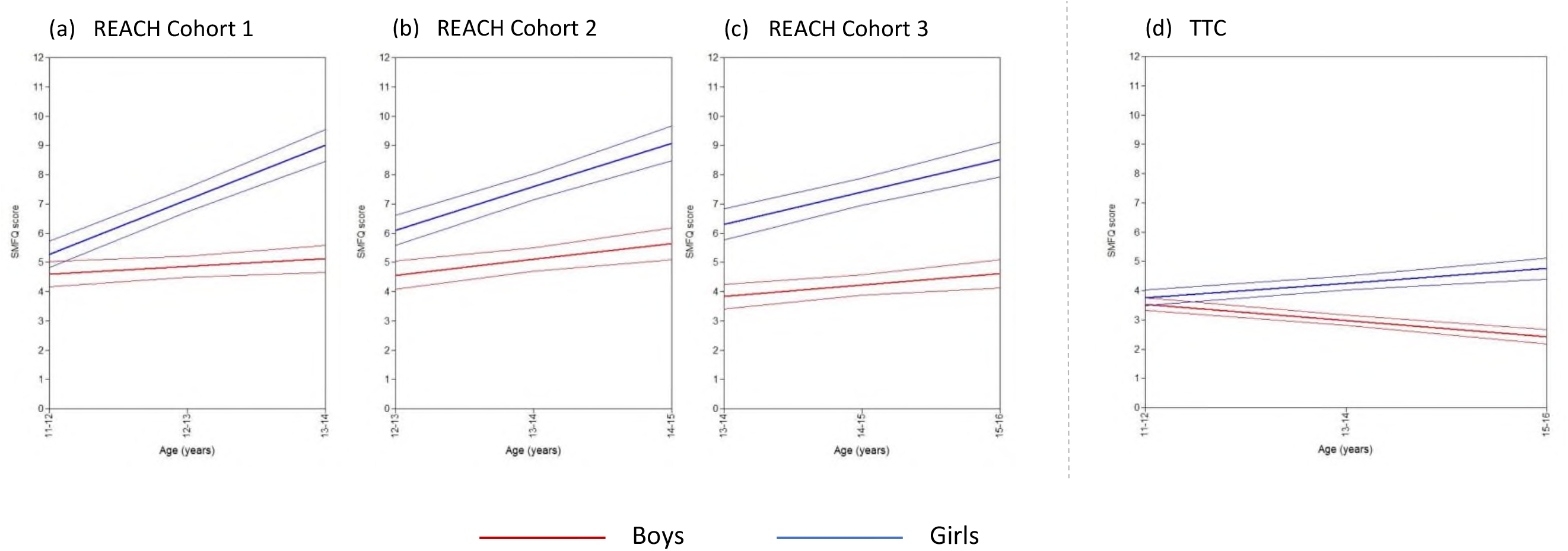
Latent growth curve models with the three REACH cohorts separated.

## REFERENCES

1. Piccinelli M, Wilkinson G. Gender differences in depression. Critical review. Br J Psychiatry 2000; 177: 486–92.

2. Kuehner C. Why is depression more common among women than among men? Lancet Psychiatry 2017; 4(2): 146–58.

3. Knowles G, Gayer-Anderson C, Beards S, et al. Mental distress among young people in inner cities: the Resilience, Ethnicity and AdolesCent Mental Health (REACH) study. J Epidemiol Community Health 2021; 75(6): 515–22.

4. Terhaag S, Fitzsimons E, Daraganova G, Patalay P. Sex, ethnic and socioeconomic inequalities and trajectories in child and adolescent mental health in Australia and the UK: findings from national prospective longitudinal studies. J Child Psychol Psychiatry 2021; 62(10): 1255–67.

5. Salk RH, Hyde JS, Abramson LY. Gender differences in depression in representative national samples: Meta-analyses of diagnoses and symptoms. Psychol Bull 2017; 143(8): 783–822.

6. Platt J, Prins S, Bates L, Keyes K. Unequal depression for equal work? How the wage gap explains gendered disparities in mood disorders. Soc Sci Med 2016; 149: 1–8.

7. Van de Velde S, Bracke P, Levecque K. Gender differences in depression in 23 European countries. Cross-national variation in the gender gap in depression. Soc Sci Med 2010; 71(2): 305–13.

8. Bentivegna F, Patalay P. The impact of sexual violence in mid-adolescence on mental health: a UK population-based longitudinal study. Lancet Psychiatry 2022; 9(11): 874–83.

9. Campbell OLK, Bann D, Patalay P. The gender gap in adolescent mental health: A cross-national investigation of 566,829 adolescents across 73 countries. SSM Popul Health 2021; 13: 100742.

10. Houri D, Nam EW, Choe EH, Min LZ, Matsumoto K. The mental health of adolescent school children: a comparison among Japan, Korea, and China. Global Health Promotion 2012; 19(3): 32–41.

11. Wade TJ, Cairney J, Pevalin DJ. Emergence of gender differences in depression during adolescence: national panel results from three countries. J Am Acad Child Adolesc Psychiatry 2002; 41(2): 190–8.

12. Lewis AJ, Rowland B, Tran A, et al. Adolescent depressive symptoms in India, Australia and USA: Exploratory Structural Equation Modelling of cross-national invariance and predictions by gender and age. J Affect Disord 2017; 212: 150–9.

13. Ando S, Nishida A, Yamasaki S, et al. Cohort Profile: The Tokyo Teen Cohort study (TTC). Int J Epidemiol 2019; 48(5): 1414-g.

14. Knowles G, Gayer-Anderson C, Blakey R, et al. Cohort Profile: Resilience, Ethnicity and AdolesCent mental Health (REACH). Int J Epidemiol 2022; 51(5): e303–e13.

15. World Economic Forum. Global Gender Gap Report 2023. Available at https://www3.weforum.org/docs/WEF_GGGR_2023.pdf?_gl=1*u6i59r*_up*MQ..&gclid=Cj0KCQjw8NilBhDOARIsAHzpbLA_r_tQcXI-lG3KqIpKiJQa5ZZyD_1R3a3H3hei-ZLNduhGI5YoxjMaAkMbEALw_wcB. Last accessed: 31^st^ July 2023.

16. Angold A, Costello EJ, Messer SC, Pickles A. Development of a short questionnaire for use in epidemiological studies of depression in children and adolescents. International journal of methods in psychiatric research 1995.

17. Liu Y, Millsap RE, West SG, Tein JY, Tanaka R, Grimm KJ. Testing measurement invariance in longitudinal data with ordered-categorical measures. Psychol Methods 2017; 22(3): 486–506.

18. Barrett P. Structural equation modelling: Adjudging model fit. Personality and Individual Differences 2007; 42(5): 815–24.

19. Chen FF. Sensitivity of Goodness of Fit Indexes to Lack of Measurement Invariance. Structural Equation Modeling: A Multidisciplinary Journal 2007; 14(3): 464–504.

20. McElroy E, Villadsen A, Patalay P, et al. Harmonisation and Measurement Properties of Mental Health Measures in Six British Cohorts. London, UK. CLOSER, 2020. Available at https://www.closer.ac.uk/wp-content/uploads/210715-Harmonisation-measurement-properties-mental-health-measures-british-cohorts.pdf. Last Accessed: 7^th^ August 2023.

201. Norman H, Elliot M. Measuring Paternal Involvement in Childcare and Housework. Sociological Research Online 2015; 20(2): 40–57.

22. Organisation for Economic Co-operation and Development. Violence against women (indicator). doi: 10.1787/f1eb4876-en. Accessed 30 March 2023 (2019 statistics); 24 July 2023 (2023 statistics).

23. Brimblecombe N, Knapp M, King D, Stevens M, Cartagena Farias J. The high cost of unpaid care by young people: health and economic impacts of providing unpaid care. BMC Public Health 2020; 20(1): 1115.

24. Balzer BWR, Duke S-A, Hawke CI, Steinbeck KS. The effects of estradiol on mood and behavior in human female adolescents: a systematic review. European Journal of Pediatrics 2015; 174(3): 289–98.

25. Skoog T, Bayram Özdemir S, Stattin H. Understanding the Link Between Pubertal Timing in Girls and the Development of Depressive Symptoms: The Role of Sexual Harassment. J Youth Adolesc. 2016;45(2):316–327.

26. Stentiford L, Koutsouris G, Allan A. Girls, mental health and academic achievement: a qualitative systematic review. Educational Review 2021: 1–31.

27. The Economist Intelligence Unit. Safe Cities Index 2021. Available at https://safecities.economist.com/safe-cities-2021-whitepaper/. Last accessed: 24th July 2023.

28. Blakey R, Morgan C, Gayer-Anderson C, et al. Prevalence of conduct problems and social risk factors in ethnically diverse inner-city schools. BMC Public Health 2021; 21(1): 849.

29. Wickham S, Whitehead M, Taylor-Robinson D, Barr B. The effect of a transition into poverty on child and maternal mental health: a longitudinal analysis of the UK Millennium Cohort Study. Lancet Public Health 2017; 2(3): e141–e8.

30. Cybulski L, Ashcroft DM, Carr MJ, et al. Temporal trends in annual incidence rates for psychiatric disorders and self-harm among children and adolescents in the UK, 2003-2018. BMC Psychiatry 2021; 21(1): 229.

